# Effectiveness of NVX-CoV2373 and BNT162b2 COVID-19 Vaccination in South Korean Adolescents

**DOI:** 10.1101/2025.04.11.25325672

**Authors:** Eunseon Gwak, Seung-Ah Choe, Kyuwon Kim, Erdenetuya Bolormaa, Manuela H. Gschwend, Jonathan Fix, Muruga Vadivale, Matthew D. Rousculp, Young June Choe

## Abstract

**Purpose:** Adolescents can have severe/chronic outcomes from COVID-19. Real-word data on relative vaccine effectiveness (rVE) between mRNA- and protein-based vaccines are limited, and more data are needed on disease outcomes in this age group.

**Methods:** The K-COV-N database, COVID-19 vaccine registry, and health insurance claims were retrospectively reviewed to identify adolescents (12–18-year-olds) in South Korea who received a homologous primary series of NVX-CoV2373 or BNT162b2 and a heterologous or homologous third vaccine dose. Vaccine recipients were propensity score matched to reduce confounding baseline factors. Adjusted hazard ratios (aHRs) for any medically attended COVID- 19 post vaccination (starting 14 days post primary series and 7 days post third dose) were calculated to assess rVE every 30 days through a 180-day risk window.

**Results:** From February to December 2022, 3174 and 6253 doses of NVX-CoV2373 and BNT162b2, respectively, were administered to South Korean adolescents. Individuals who received NVX-CoV2373 tended to be older, have a disability, and/or have a prior SARS-CoV-2 infection. Propensity score matching resulted in 107 individuals in each primary series group and 701 and 1417 individuals in the NVX-CoV2373 and BNT162b2 third-dose groups, respectively. The aHR (95% CI) for NVX-CoV2373 compared with BNT162b2 for medically attended COVID-19 in the 180-day risk window was 0.57 (0.31–1.05) for the primary series and 0.68 (0.54–0.84) for the third dose.

**Discussion:** These results suggest that NVX-CoV2373 may provide more robust protection against medically attended COVID-19, compared to BNT162b2, both as a homologous primary series and as a homologous or heterologous third dose.

**Implications and Contributions:** This study provides essential data on the real-world effectiveness of a protein-based (NVX- CoV2373) and an mRNA-based (BNT162b2) COVID-19 vaccine in adolescents. A homologous primary series and homologous/heterologous third dose of NVX-CoV2373 provided more robust protection than BNT162b2 at preventing medically attended COVID-19, with protection demonstrated through 6 months post vaccination.

## Introduction

COVID-19 remains a significant global health challenge, complicated by the emergence of SARS-CoV-2 variants that require regular updates to the target spike protein(s) included in these vaccines [1]. In South Korea and across the world, various COVID-19 vaccine formulations have been made available for adolescents, including the mRNA-based Pfizer– BioNTech COVID-19 Vaccine and the protein-based Novavax COVID-19 Vaccine [2,3]. While clinical trials of these vaccines have demonstrated safety and efficacy in adolescents [4–8], real-world data on their comparative effectiveness in this population are limited. One study in the United States, assessing online data of COVID-19 cases from the start of the pandemic up to December 2020 (prior to vaccine availability), reported a statistically higher incidence rate in adolescents (aged 10–19 years), in comparison with adults (aged ≥60 years), in 10 of 13 states [9]. Collecting information in larger populations and other regions and including data on infection and disease rates based on type of vaccination (eg, protein vs mRNA and dose) are particularly crucial in the context of evolving viral variants and the unique immune responses of this age group.

The World Health Organization and Korean Disease Control and Prevention Agency have authorized receipt of a COVID-19 vaccine for adolescents, with additional doses suggested for those who are immunocompromised [10,11]. Immunization campaigns continue to have an impact on COVID-19–related mortality in the adolescent population; however, the implementation and success of these vary by region. For example, Japan has a universal vaccination policy and higher immunization rates than does South Korea, which has a risk-based vaccination policy [12]. Notably, a comparative analysis of COVID-19–related mortality among those aged 10–19 years within these regions demonstrated a >2.5-fold decrease in deaths attributed to COVID-19 in Japan versus South Korea [12]. Additionally, severe illness/complications (eg, multisystem inflammatory syndrome in children) and post-COVID conditions (PCC) can occur in a small proportion of infected children [13], and vaccination can significantly reduce the risk of these occurring [14,15]. This risk reduction is critical, as the development of chronic symptoms, such as PCC, not only affects health but could also negatively affect academic performance and social development [16]. For example, one study highlighted the importance of school attendance, reporting that school absence, specifically due to illness in 16- to 18-year-olds, was significantly associated with an increased likelihood of not being in education, employment, or training after secondary education [17]. Notably, in children aged 5–17 years hospitalized with COVID-19, correlations have been described for reduced school absence if they were previously vaccinated, in comparison with an unvaccinated comparator group.

Although adolescents generally experience milder COVID-19 symptoms than adults [18], their infection can result in an index case for transmitting infection to more vulnerable populations. In a South African study that included waves of early SARS-CoV-2 variants (ie, alpha [B.1.1.7], beta [B.1.351], delta [B.1.617.2]) [19], household transmission increased in each successive wave, as well as proportionately more infections among adolescents [20]. In that study, most overall infections were found to be asymptomatic, with lack of symptoms being most common in those aged younger than 19 years. Notably, transmission of SARS-CoV-2 was similar whether from symptomatic or asymptomatic individuals.

Lee et al. conducted a retrospective assessment of relative vaccine effectiveness (rVE) of a primary series of the NVX-CoV2373 and BNT162b2 COVID-19 vaccines in adolescents (aged 12–17 years) in South Korea [21]. Results indicated that a primary series of these vaccines were considered noninferior to each other for SARS-CoV-2 infection over a course of 13 weeks among the two matched cohorts (each n = 465).

This study seeks to fill gaps in real-world vaccine effectiveness (VE) data by investigating the rVE of NVX-CoV2373 in comparison with BNT162b2 in preventing medically attended COVID-19 in South Korean adolescents during the period of Omicron variant predominance. Importantly, the data described here include follow-up through 6 months post vaccination as well as cohorts completing a homologous primary series and homologous or heterologous third vaccine dose. Through population-based data, valuable insights into the real-world performance of these vaccines are provided and can inform public health strategies and vaccination policies.

## Methods

### Study population and data acquisition

This study utilized data from the Korea Disease Control and Prevention Agency– COVID-19–National Health Insurance Service (K-COV-N) cohort database, a comprehensive resource integrating COVID-19 vaccination records, SARS-CoV-2 infection surveillance data, and health insurance claims. Adolescents aged 12–18 years who received either NVX-CoV2373 or BNT162b2 in South Korea between February 1 and December 31, 2022, and had at least 1 year of health data available prior to vaccination, were the focus of this study. To compare VE, this study grouped the identified vaccine recipients by vaccination status into a primary series group (ie, received a homologous primary series of NVX-CoV2373 or BNT162b2) and a third-dose group (ie, received a homologous or heterologous dose of NVX-CoV2373 or BNT162b2 after a homologous primary series of either COVID-19 vaccine).

### Outcomes

The primary outcome of interest was medically attended COVID-19. Data were collected from outpatient, emergency department, and intensive care unit records. Specific treatment codes for admission to an intensive care unit, coupled with a primary or secondary diagnosis of conditions related to SARS-CoV-2/COVID-19 (ie, International Classification of Diseases 10th rev. codes B342, B972, Z208, Z290, Z115, U181, Z038, U071, U072, U08, U09, and U10), were used to identify medically attended COVID-19.

### Statistical analysis

Propensity score matching was used to ensure balanced cohorts for appropriate comparison between individuals who received either NVX-CoV2373 or BNT162b2. Propensity scores were estimated on the basis of demographic and clinical factors, including income level, residential region (inside or outside of the Seoul capital area), disability status, Charlson Comorbidity Index, prior SARS-CoV-2 infection history, comorbidities, and month of vaccination. The matching process paired the two groups on the basis of similar propensity scores, using a caliper of 0.25 and exact matching on the month of the vaccination. Descriptive statistics were used to assess baseline characteristics of vaccine recipients. Between-group baseline characteristics were compared using the absolute standardized difference (aSD), where aSD >0.1 indicated a significant imbalance [22].

The primary outcome measure was the rVE of NVX-CoV2373, compared with BNT162b2, in preventing medically attended COVID-19. To assess rVE, crude incidence rates (IRs) per 1000 person-days and adjusted hazard ratios (aHRs), which considered time-varying vaccination status, were calculated. The Cox proportional-hazards model was used to calculate the aHRs of medically attended COVID-19 for NVX-CoV2373 in comparison with BNT162b2.

The index date was the day of vaccination, with follow-up beginning 14 days post primary series and 7 days post third dose, and with data collection occurring for up to 180 days or until the earliest of a COVID-19 outcome, next dose administration, or the end of the study period.

Outcomes were evaluated at the following 30-day risk windows from the index date: 30, 60, 90, 120, 150, and 180 days. A negative control analysis examining the association between vaccination and medically attended COVID-19 in the first 0–5 days after vaccination was performed to assess potential residual or uncontrolled confounding.

All statistical analyses were performed using SAS^®^ 9.4 software (SAS Institute, Cary, NC, USA), and the study received ethical approval from the Korea University Anam Hospital institutional review board (IRB No. 2023AN0124).

## Results

### Vaccine doses and study population

A total of 4827 NVX-CoV2373 and 41,789 BNT162b2 doses were administered to the national population aged 12–18 years (**Figure 1**). Restricting the analysis to second or higher doses, homologous primary series, and doses administered between February 1 and December 31, 2022, resulted in 3174 NVX-CoV2373 and 6253 BNT162b2 doses being included in this analysis. Because the number of fourth doses of NVX-CoV2373 (n = 25) and BNT162b2 (n = 6) was small, the fourth-dose group was excluded. After propensity score matching to balance baseline characteristics, the final analysis included 107 recipients of each vaccine in the primary series group and 701 and 1417 recipients of NVX-CoV2373 and BNT162b2 in the third-dose groups, respectively.

**Figure 1.** Study design for rVE against medically attended COVID-19 comparing NVX- CoV2373 and BNT162b2. ^a^Includes homologous primary series only. ^b^Follow-up started 14 days after the second dose of the primary series or 7 days after the third vaccine dose and continued until censoring. Risk windows for outcomes were 30, 60, 90, 120, 150, and 180 days after the start of follow-up. ^c^Propensity score matching based on demographic and clinical characteristics and month of vaccination. rVE = relative vaccine effectiveness.

Descriptive statistics before propensity score matching indicated that individuals who received NVX-CoV2373 (in comparison with BNT162b2) as a homologous primary series tended to be older (18 years of age versus <18), have a disability, and have a history of a prior SARS-CoV-2 infection (**Table S1**). For third-dose groups, NVX-CoV2073 recipients were more likely to be younger, live in the capital area, have medical comorbidities, have a disability, be in the highest income quartile, and/or have a prior SARS-CoV-2 infection. After matching, the differences were largely balanced, with minimal residual discrepancies remaining (**Table 1**).

**Table 1.** Matched baseline characteristics for primary series and third dose groups, by vaccine type.

| Variable, n (%) | Primary series |  |  | Third dose |  |  |
| --- | --- | --- | --- | --- | --- | --- |
|  | NVX-CoV2073 | BNT162b2 | aSD<sup>a</sup> | NVX-CoV2073 | BNT162b2 | aSD<sup>a</sup> |
| n | 107 | 107 |  | 701 | 1417 |  |
| Age (years) |  |  |  |  |  |  |
| 12 | 7 (6.5) | 14 (13.1) | <0.01 | 4 (0.6) | 3 (0.2) | 0.15 |
| 13 | 7 (6.5) | 2 (1.9) |  | 3 (0.4) | 20 (1.4) |  |
| 14 | 1 (0.9) | 2 (1.9) |  | 1 (0.1) | 18 (1.3) |  |
| 15 | 4 (3.7) | 1 (0.9) |  | 1 (0.1) | 45 (3.2) |  |
| 16 | 1 (0.9) | 16 (15.0) |  | 42 (6.0) | 88 (6.2) |  |
| 17 | 1 (0.9) | 8 (7.5) |  | 37 (5.3) | 185 (13.1) |  |
| 18 | 86 (80.4) | 64 (59.8) |  | 613 (87.5) | 1058 (74.7) |  |
| Sex |  |  |  |  |  |  |
| Male | 47 (43.9) | 51 (47.7) | 0.08 | 341 (48.6) | 824 (58.2) | 0.19 |
| Female | 60 (56.1) | 56 (52.3) |  | 360 (51.4) | 593 (41.9) |  |
| Non-capital area<sup>b</sup> | 53 (49.5) | 54 (50.5) | 0.02 | 271 (38.7) | 669 (47.2) | 0.17 |
| CCI ≥3 | 0 (0.0) | 0 (0.0) | NA | 1 (0.1) | 1 (0.1) | 0.02 |
| Disability | 4 (3.7) | 5 (4.7) | 0.05 | 27 (3.9) | 17 (1.2) | 0.19 |
| Parental income level<sup>c</sup> |  |  |  |  |  |  |
| Medical aid beneficiary | 1 (0.9) | 2 (1.9) | 0.06 | 38 (5.5) | 50 (3.6) | 0.10 |
| 1Q | 7 (6.6) | 12 (11.4) | 0.16 | 75 (10.8) | 145 (10.4) | 0.02 |
| 2Q | 26 (24.5) | 19 (18.1) | 0.19 | 82 (11.8) | 205 (14.6) | 0.08 |
| 3Q | 23 (21.7) | 27 (25.7) | 0.10 | 98 (14.1) | 232 (16.6) | 0.06 |
| 4Q (richest) | 49 (46.2) | 45 (42.9) | 0.08 | 401 (57.8) | 768 (54.9) | 0.06 |
| Prior SARS-CoV-2 infection | 16 (15.0) | 22 (20.6) | 0.17 | 189 (27.0) | 435 (30.7) | 0.10 |

<sup>a</sup>aSD values >0.1 indicate a significant imbalance between vaccine groups. <sup>b</sup>Indication that vaccine recipients were living outside the Seoul capital area. <sup>c</sup>Income was categorized as medical aid beneficiary (no/lowest income) and then into 1Q (10%–40%), 2Q (>40%–60%), 3Q (>60%–80%), and 4Q (>80%–100%; highest income). aSD = absolute standardized difference; CCI = Charlson comorbidity index; NA = not applicable; Q = quartile.

### Medically attended COVID-19

Among the primary series recipients, the IR/1000 person-months (95% CI) of medically attended COVID-19 was 1.31 (0.86–1.99) in the NVX-CoV2373 group (22 cases) and 1.52 (1.02–2.27) in the BNT162b2 group (24 cases) during the 180-day risk window, resulting in an aHR of 0.57 (95% CI: 0.31–1.05); **Table 2**; **Figure 2**). IR/1000 person-months (95% CI) in the third-dose groups for NVX-CoV2373 recipients (112 cases) was 0.96 (0.79–1.15) and for BNT162b2 recipients (305 cases) was 1.32 (1.18–1.48), with a resulting aHR of 0.68 (95% CI: 0.54–0.84). During the study period, there was one case of severe disease following a third dose of NVX-CoV2373; there were no severe cases following a third dose of BNT162b2.

**Figure 2.**
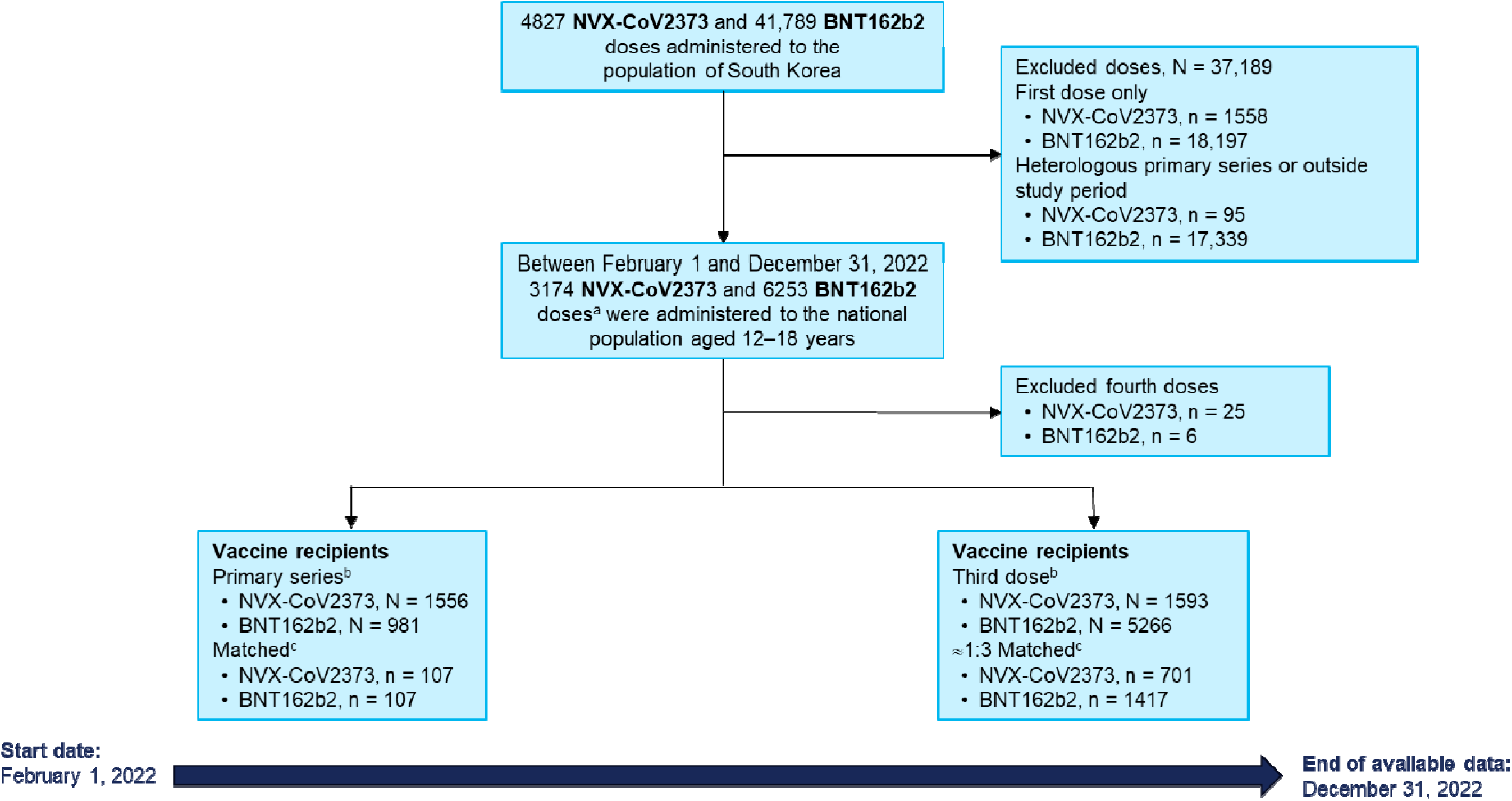
Study design for rVE against medically attended COVID-19 comparing NVX-CoV2373 and BNT162b2

**Figure 3.**
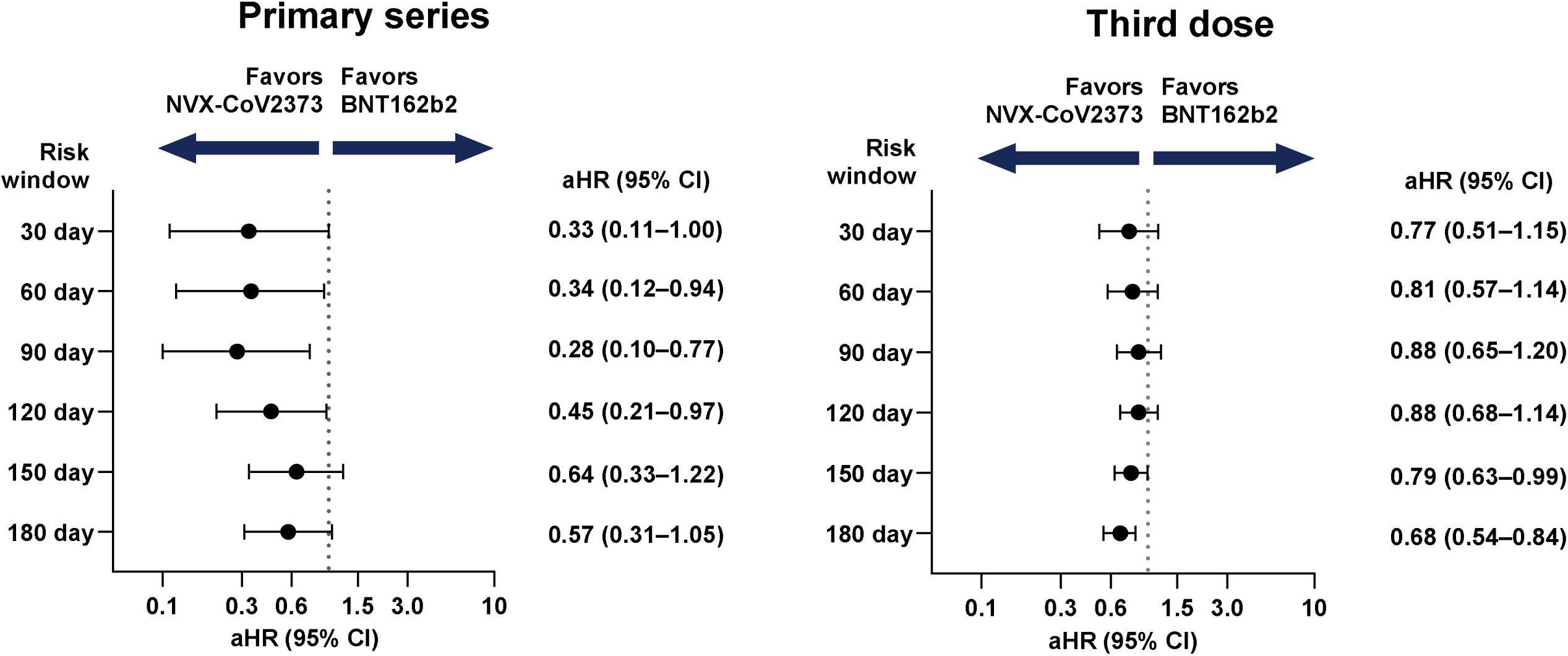
Risk estimates (aHRs) for medically attended COVID-19 after a homologous primary series (left) and homologous or heterologous third dose (right) of NVX-CoV2373 or BNT162b2.

**Table 2.**
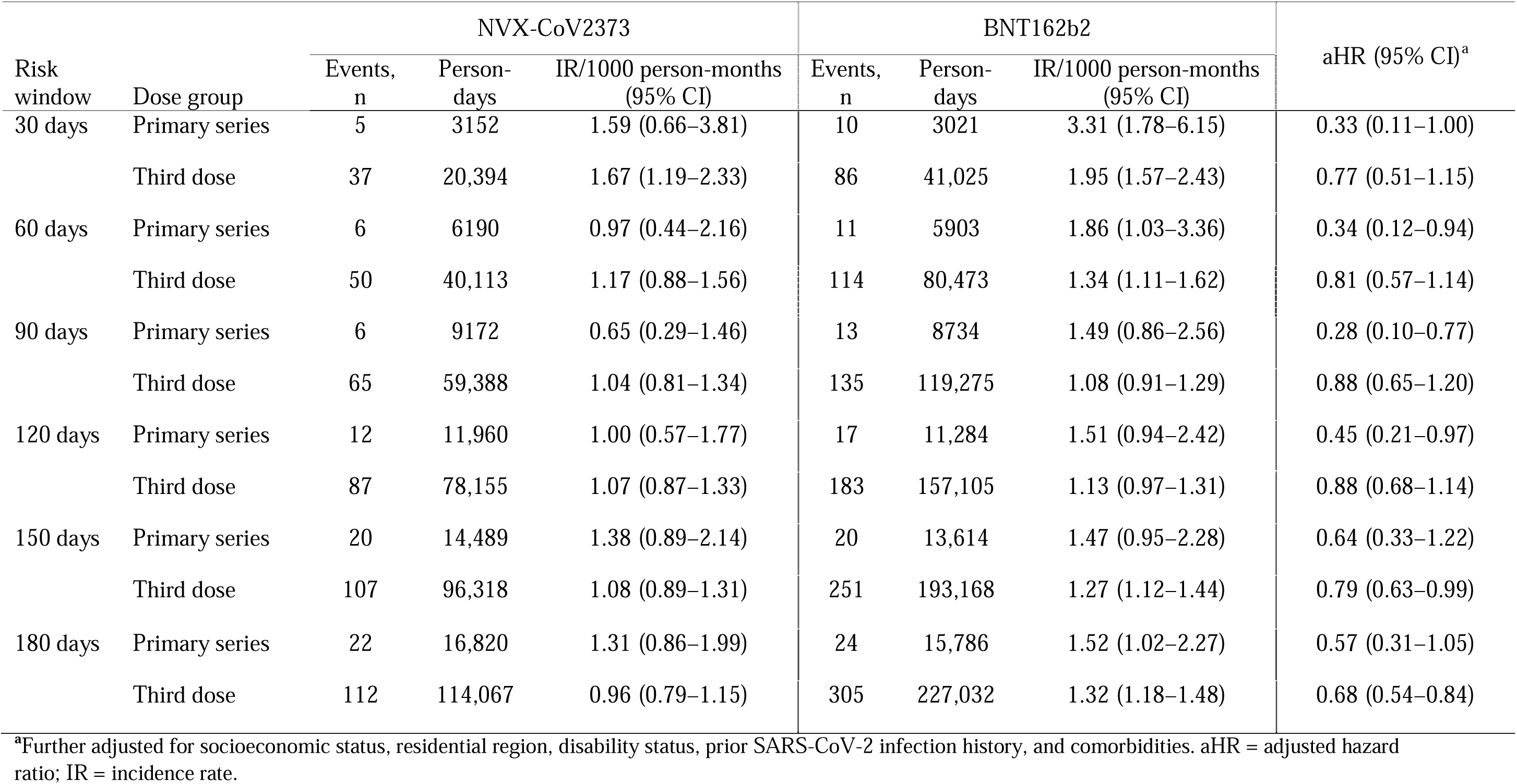
The IR and aHR of medically attended COVID-19 after receipt of a homologous primary series and homologous or heterologous third vaccine dose^a^ of NVX-CoV2373 or BNT162b2 (stratified by risk window since vaccination)

| Risk window | Dose group | NVX-CoV2373 |  |  | BNT162b2 |  |  | aHR (95% CI)<sup>a</sup> |
| --- | --- | --- | --- | --- | --- | --- | --- | --- |
|  |  | Events, n | Person-days | IR/1000 person-months (95% CI) | Events, n | Person-days | IR/1000 person-months (95% CI) |  |
| 30 days | Primary series | 5 | 3152 | 1.59 (0.66–3.81) | 10 | 3021 | 3.31 (1.78–6.15) | 0.33 (0.11–1.00) |
|  | Third dose | 37 | 20,394 | 1.67 (1.19–2.33) | 86 | 41,025 | 1.95 (1.57–2.43) | 0.77 (0.51–1.15) |
| 60 days | Primary series | 6 | 6190 | 0.97 (0.44–2.16) | 11 | 5903 | 1.86 (1.03–3.36) | 0.34 (0.12–0.94) |
|  | Third dose | 50 | 40,113 | 1.17 (0.88–1.56) | 114 | 80,473 | 1.34 (1.11–1.62) | 0.81 (0.57–1.14) |
| 90 days | Primary series | 6 | 9172 | 0.65 (0.29–1.46) | 13 | 8734 | 1.49 (0.86–2.56) | 0.28 (0.10–0.77) |
|  | Third dose | 65 | 59,388 | 1.04 (0.81–1.34) | 135 | 119,275 | 1.08 (0.91–1.29) | 0.88 (0.65–1.20) |
| 120 days | Primary series | 12 | 11,960 | 1.00 (0.57–1.77) | 17 | 11,284 | 1.51 (0.94–2.42) | 0.45 (0.21–0.97) |
|  | Third dose | 87 | 78,155 | 1.07 (0.87–1.33) | 183 | 157,105 | 1.13 (0.97–1.31) | 0.88 (0.68–1.14) |
| 150 days | Primary series | 20 | 14,489 | 1.38 (0.89–2.14) | 20 | 13,614 | 1.47 (0.95–2.28) | 0.64 (0.33–1.22) |
|  | Third dose | 107 | 96,318 | 1.08 (0.89–1.31) | 251 | 193,168 | 1.27 (1.12–1.44) | 0.79 (0.63–0.99) |
| 180 days | Primary series | 22 | 16,820 | 1.31 (0.86–1.99) | 24 | 15,786 | 1.52 (1.02–2.27) | 0.57 (0.31–1.05) |
|  | Third dose | 112 | 114,067 | 0.96 (0.79–1.15) | 305 | 227,032 | 1.32 (1.18–1.48) | 0.68 (0.54–0.84) |

<sup>a</sup>Further adjusted for socioeconomic status, residential region, disability status, prior SARS-CoV-2 infection history, and comorbidities. aHR = adjusted hazard ratio; IR = incidence rate.

### Negative control analysis

In the matched pairs, the odds of all infections during days 0−5 post third dose did not significantly differ between the NVX-CoV2373 and BNT162b2 groups (adjusted odds ratio = 1.10, 95% CI: 0.46–2.60; **Table 3**).

**Table 3.** The frequency and aOR of medically attended COVID-19 comparing NVX-CoV2373 and BNT162b2 recipients within 0–5 days post vaccination.

|  | NVX-CoV2373 |  |  | BNT162b2 |  |  | *P* | aOR (95% CI) |
| --- | --- | --- | --- | --- | --- | --- | --- | --- |
|  | Recipients, n | Events, n | % | Recipients, n | Events, n | % |  |  |
| Primary series | 107 | 2 | 1.87 | 107 | 0 | 0 | 0.156 |  |
| Third dose | 701 | 8 | 1.14 | 1417 | 20 | 1.41 | 0.609 | 1.10 (0.46–2.60) |

aOR = adjusted odds ratio.

## Discussion

In this study, the rVE of NVX-CoV2373 compared to BNT162b2 in preventing medically attended COVID-19 in South Korean adolescents during the period of Omicron variant predominance (eg, BA.1, BA.4/5, XBB) [23] was investigated. These findings suggest that NVX-CoV2373 offered additional protection from medically attended disease, in comparison with BNT162b2, in this population. Additionally, risk estimates (aHR [95% CI]) observed through 6 months post vaccination suggest continued protection after a homologous primary series (0.57 [0.31–1.05]) or third dose (0.68 [0.54–0.84]) of NVX-CoV2373.

The effectiveness of COVID-19 vaccines in adolescents is crucial for tempering illness in this population, as well as reducing potential transmission to other populations. For example, in a meta-analysis of SARS-CoV-2 transmission studies involving children aged ≤18 years, there was a child-to-adult transmission rate of 47% [24]. Studies have shown that primary COVID-19 vaccine series (both protein and mRNA based) and subsequent doses are highly effective in preventing infection, hospitalization, and death in adolescents [4–8]. A key factor in immune defense against invading pathogens is protection at viral entry points of the respiratory tract (eg, nose and mouth) and the subsequent mucosal immune response. While mucosal immunoglobulin A (IgA) levels increase readily with vaccination, these wane rapidly [25], and repeat dosing with mRNA-based vaccines may not reestablish IgA to protective levels [26]. In a preliminary assessment, anti-spike IgA responses were observed against various SARS-CoV-2 strains with Novavax COVID-19 vaccination (targeting XBB.1.5), after ≥2 doses of an mRNA-based COVID-19 vaccine [27].

These findings are consistent with a previous comparative randomized controlled trial in adolescents [28]. The identified differences in rVE could be attributed to multiple factors, including the different components and mechanisms of action of the two vaccines. For example, the Novavax COVID-19 Vaccine contains an already translated recombinant spike protein (instead of mRNA) and Matrix-M™ adjuvant, which can be a potent immune activator [29] and potentially involved in the generation of cross-reactive CD4 T-cells [30]. Notably, NVX-CoV2373 elicited a SARS-CoV-2–specific immune response in people with multiple sclerosis who had insufficient immune responses to previous mRNA/viral vector vaccination [31].

Clinical and real-world comparative analyses of the diverse types of vaccines are limited.

The phase 2 Com-COV3 study in the United Kingdom investigated outcomes in adolescents (aged 12–16 years) who received a second COVID-19 vaccination of BNT162b2 (30 µg or 10 µg) or NVX-CoV2373 during the study after having received an initial dose of BNT162b2 [28]. At 28 days post second dose, anti-immunoglobulin G levels were comparable between the BNT162b2 30 µg and NVX-CoV2373 groups; however, higher 28-day neutralizing antibody titers and 14-day cellular immune responses were observed with NVX-CoV2373 than with either of the BNT162b2 groups. Additionally, the NVX-CoV2373 group had the lowest proportion of breakthrough infections through 236 days (≈8 months) post study vaccination. In the rVE study for SARS-CoV-2 infection in adolescents by Lee et al., the risk estimate for infection after a primary series of NVX-CoV2373 versus BNT162b2 was 1.46 (95% CI: 0.68–3.22) at 13 weeks post vaccination [21]. This assessment of SARS-CoV-2 infection has differences from the rVE data of medically attended COVID-19 described here. For example, the differences in unmatched populations from the analysis described in the Results section highlight the importance of propensity score matching in mitigating potential confounding factors when assessing rVE. Key limitations to consider from the study by Lee et al. are censoring patients at 7 weeks, skewed age-group distribution, and lack of variables for comprehensive matching.

This study has several limitations that merit consideration. Adolescents who received a primary series after February 2022 may have restricted eligibility and generalizability, as most individuals had likely received both initial doses of BNT162b2 prior to this date. Additionally, focusing the analysis to NVX-CoV2373 and BNT162b2 homologous primary series and homologous/heterologous third doses may pose some restrictions in generalizing findings to the broader adolescent population. Control for confounding factors was performed through propensity score matching and evaluated through assessing medically attended COVID-19 0–5 days post vaccination; however, the possibility of residual confounding factors or the introduction of potential biases based on the observational nature of the study cannot be completely ruled out. For example, reports of myocarditis after receipt of an mRNA-based COVID-19 vaccine [32] may create a bias for parental choice of vaccine receipt and/or formulation (mRNA or protein) selection. Another limitation may be the relatively small sample size for the overall analysis and restricted assessment of rVE of severe COVID-19, because there was only one severe case throughout the 6-month follow-up; although, this is not unexpected on the basis of lower severity of illness typical in adolescents versus adults [18]. Finally, the study period coincided with the predominance of Omicron variants [23]; therefore, findings may not be generalizable to periods with different dominant variants.

In conclusion, this study provides valuable insights into the comparative effectiveness of NVX-CoV2373 and BNT162b2 in preventing medically attended COVID-19 among South Korean adolescents, suggesting that NVX-CoV2373 may offer additional protection in comparison with BNT162b2 in this population. Evaluation of doses beyond the primary series and the long-term effectiveness of the vaccines are key considerations for future research.

Overall, these findings have important implications for public health strategies and vaccination policies, particularly in the context of evolving viral variants and the unique immune responses of adolescents.

## Funding

Novavax Inc.

## Data Availability

The data used in this study are from the K-CoV-N cohort, which is not publicly available. Researchers can apply for access to these data through the National Health Insurance Service (NHIS) and Korea Disease Control and Prevention (KDCA) data linkage system. Applications can be submitted via the Health Care Data Linkage (HCDL) website (https://hcdl.mohw.go.kr/) and are subject to review by the relevant authorities.

## Acknowledgments

Medical writing and editorial support for the development of this manuscript, under the direction of the authors, were provided by Kelly M. Fahrbach, Ph.D., CMPP, and Celia Nelson of Ashfield MedComms (US), an Inizio company, and was funded by Novavax, Inc. This study used the Korea Disease Control and Prevention Agency (KDCA) and National Health Insurance Service (NHIS) databases for policy and academic research (KDCA-NHIS-2023-1-494). The conclusions of this study are not related to this institution.

## Conflicts of Interest

E.G., S.-A.C., K.K., E.B., and Y.J.C. are study investigators and do not have any conflicts of interest to report. M.H.G., J.F., and M.D.R. are salaried employees of and may own stock in Novavax, Inc. M.V. was a salaried employee of Novavax, Inc., and may own stock.

## Author Contributions

E.G., S.-A.C., M.V., M.H.G., M.D.R., and Y.J.C. conceptualized the trial and contributed to its design. E.G., S.-A.C., K.K., and E.B. were involved in the extraction and analysis of data. E.G., S.-A.C., K.K., E.B., J.F., M.V., M.H.G., M.D.R., and Y.J.C. were involved in the interpretation of data. E.G., S.-A.C., K.K., E.B., and Y.J.C. verified the data. E.G., S.-A.C., K.K., E.B., J.F., M.V., M.H.G., M.D.R., and Y.J.C. drafted the manuscript. E.G., S.-A.C., and K.K. performed the statistical analyses. S.-A.C., J.F., M.H.G., M.D.R., and Y.J.C. supervised the study. All authors critically reviewed and approved the manuscript.

## Abbreviations

aHR, adjusted hazard ratio; aSD, absolute standardized difference; IgA, immunoglobulin A; IR, incidence rate; rVE, relative vaccine effectiveness.

---

**Table S1.**
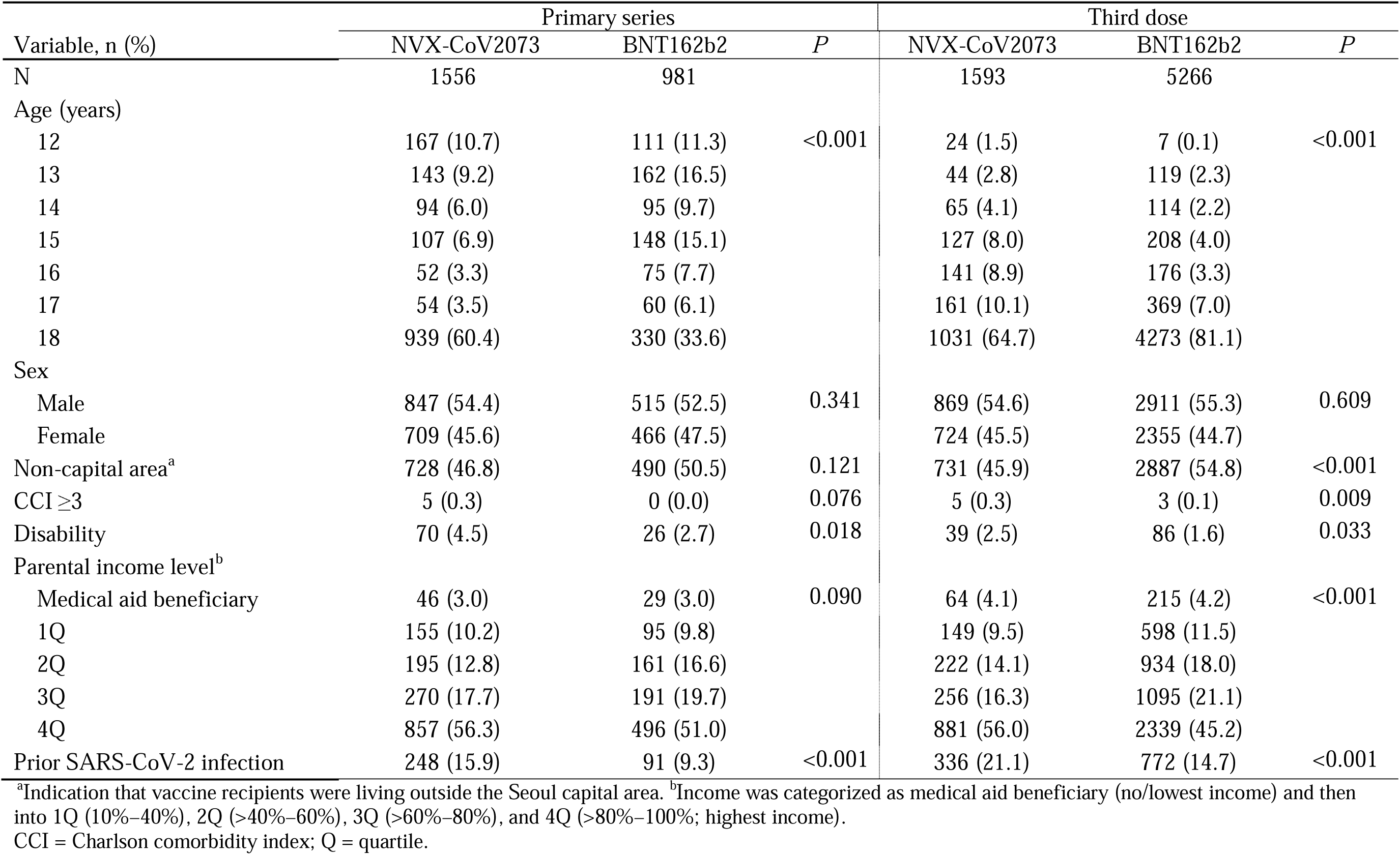
Baseline characteristics of vaccine recipients, by dose group, before propensity score matching.

| Variable, n (%) | Primary series |  |  | Third dose |  |  |
| --- | --- | --- | --- | --- | --- | --- |
|  | NVX-CoV2073 | BNT162b2 | *P* | NVX-CoV2073 | BNT162b2 | *P* |
| N | 1556 | 981 |  | 1593 | 5266 |  |
| Age (years) |  |  |  |  |  |  |
| 12 | 167 (10.7) | 111 (11.3) | <0.001 | 24 (1.5) | 7 (0.1) | <0.001 |
| 13 | 143 (9.2) | 162 (16.5) |  | 44 (2.8) | 119 (2.3) |  |
| 14 | 94 (6.0) | 95 (9.7) |  | 65 (4.1) | 114 (2.2) |  |
| 15 | 107 (6.9) | 148 (15.1) |  | 127 (8.0) | 208 (4.0) |  |
| 16 | 52 (3.3) | 75 (7.7) |  | 141 (8.9) | 176 (3.3) |  |
| 17 | 54 (3.5) | 60 (6.1) |  | 161 (10.1) | 369 (7.0) |  |
| 18 | 939 (60.4) | 330 (33.6) |  | 1031 (64.7) | 4273 (81.1) |  |
| Sex |  |  |  |  |  |  |
| Male | 847 (54.4) | 515 (52.5) | 0.341 | 869 (54.6) | 2911 (55.3) | 0.609 |
| Female | 709 (45.6) | 466 (47.5) |  | 724 (45.5) | 2355 (44.7) |  |
| Non-capital area<sup>a</sup> | 728 (46.8) | 490 (50.5) | 0.121 | 731 (45.9) | 2887 (54.8) | <0.001 |
| CCI ≥3 | 5 (0.3) | 0 (0.0) | 0.076 | 5 (0.3) | 3 (0.1) | 0.009 |
| Disability | 70 (4.5) | 26 (2.7) | 0.018 | 39 (2.5) | 86 (1.6) | 0.033 |
| Parental income level<sup>b</sup> |  |  |  |  |  |  |
| Medical aid beneficiary | 46 (3.0) | 29 (3.0) | 0.090 | 64 (4.1) | 215 (4.2) | <0.001 |
| 1Q | 155 (10.2) | 95 (9.8) |  | 149 (9.5) | 598 (11.5) |  |
| 2Q | 195 (12.8) | 161 (16.6) |  | 222 (14.1) | 934 (18.0) |  |
| 3Q | 270 (17.7) | 191 (19.7) |  | 256 (16.3) | 1095 (21.1) |  |
| 4Q | 857 (56.3) | 496 (51.0) |  | 881 (56.0) | 2339 (45.2) |  |
| Prior SARS-CoV-2 infection | 248 (15.9) | 91 (9.3) | <0.001 | 336 (21.1) | 772 (14.7) | <0.001 |

<sup>a</sup>Indication that vaccine recipients were living outside the Seoul capital area. <sup>b</sup>Income was categorized as medical aid beneficiary (no/lowest income) and then into 1Q (10%–40%), 2Q (>40%–60%), 3Q (>60%–80%), and 4Q (>80%–100%; highest income). CCI = Charlson comorbidity index; Q = quartile.

## Notes

### Funding Statement

The study was funded by Novavax, Inc.

### Author Declarations

The study received ethical approval from the Korea University Anam Hospital institutional review board (IRB No. 2023AN0124).

